# Healthcare workers’ compliance with COVID-19 preventive and control measures at De Martino Hospital, Mogadishu, Somalia

**DOI:** 10.1101/2024.03.29.24305060

**Authors:** Abdullahi Ibrahim Janay, Bulent Kilic, Belgin Unal

## Abstract

**Background and objectives:** Healthcare workers are a high-risk group for COVID-19 and protecting them is crucial for healthcare delivery. Limited studies have explored compliance with infection prevention and control (IPC) practices among Somali healthcare workers. This study aimed to determine compliance with IPC practices among healthcare workers in De Martino Public Hospital, Somalia.

**Materials and methods:** A cross-sectional study was conducted at De Martino Public Hospital, Mogadishu, Somalia from August to October 2022, with the participation of 204 healthcare workers (response rate = %97). Compliance was assessed using responses to 25 questions on a five-point Likert-type scale, and the median score of 20 was used to dichotomize compliance scores. A chi-square test and logistic regression analysis were performed to check the associations between healthcare workers’ sociodemographic information, related factors to IPC, work conditions and practices on COVID-19, and IPC compliance during healthcare interventions using SPSS 23 version.

**Results:** In total, 58.3% of the participants had good compliance with IPC. There were significant associations between IPC compliance and the type of healthcare worker (doctors and doctor assistants: 72.3%, nurses and paramedical staff: 67.3%, non-clinical staff: 5.7%, p<0.01). After adjusting for potential confounding factors, compared to non-clinical staff, doctors and doctor assistants (OR: 12.11, 95% CI: 2.23-65.84) nurses and paramedical staff (OR: 21.38, 95% CI: 4.23 - 108.01) had higher compliance with infection prevention and control measures. There were no significant associations between compliance and gender, marital status, vaccination status, or smoking (p>0.05 for all).

**Conclusions:** Inadequate compliance with COVID-19 IPC measures was observed among hospital workers. Prioritizing awareness campaigns and behavior change interventions, especially among non-clinical staff, is crucial for effective COVID-19 infection prevention and control within the hospital.

## Introduction

The COVID-19 pandemic was the most recent and the worst pandemic in the 21st century, and has resulted in 765 million confirmed cases and 6.9 million deaths worldwide as of May 3, 2023 [1].

This highly infectious virus can be transmitted by asymptomatic carriers and those in the incubation period, making isolation until testing negative crucial [2]. Quarantine measures are essential for persons from endemic areas or with confirmed contact and for mild cases not requiring medical attention [3]. All age groups are susceptible to the virus, but elderly persons with underlying health conditions are at higher risk of severe illness [4]. Healthcare workers (HCWs) are also vulnerable, emphasizing the importance of infection prevention and control (IPC) practices and personal protective equipment (PPE) use [5].

IPC strategies include non-pharmaceutical interventions like school and workplace closures, event bans, stay-at-home orders, and movement restrictions, which have reduced transmission [6]. Personal protective measures are also an important component of COVİD-19 infection prevention and control [7]. Vaccines have been developed, but their effectiveness varies by variant, with better protection against severe outcomes. Vaccine effectiveness against symptomatic disease was higher for the delta variant than for the omicron variant. With previous variants, vaccine effectiveness against severe disease, including hospitalization and death, has been higher and retained for longer than effectiveness against mild disease [8].

World Health Organization (WHO) declared COVID-19 is an emergency public health problem of international concern that poses a high risk to countries with vulnerable health systems on January 30, 2020 [9]. Despite the Director General of the WHO declared the end of COVID-19 as a public health emergency and is no longer a global threat on May 5th 2023, he said that COVID-19 is still killing and warned of the risks from the new emerging variants [10]. In fragile settings like Somalia where the number of HCWs is significantly lower than required for Sustainable Development Goals, the protection of health workers is vital [11]. To protect HCWs, Enough supplies of PPE should be ensured and HCWs should be trained in using it. The environmental hygiene of hospitals and the personal hygiene of HCWs should be maintained [2]. HCWs should maintain contact reduction to reduce the number of infections [7].

It was reported from China that the potential risk of COVID-19 has largely improved the IPC behaviors of HCWs working in hospitals [12]. Determination of compliance of the hospital workers with the COVID-19 IPC practices and the factors affecting this compliance is important for the protection of hospital workers during the pandemic. Studies assessing COVID-19 IPC practices in Somali hospitals and HCWs are limited. This cross-sectional study assessed the COVID-19 IPC compliance among HCWs in De Martino Public Hospital during healthcare interventions and determined the factors affecting their compliance to improve hospital workers’ compliance with COVID-19 IPC practices.

## Materials and methods

This hospital-based cross-sectional study was performed at De Martino Public Hospital in Mogadishu, Somalia’s capital. It was built during Italy’s colonial era and is currently under the management of the Ministry of Health and Human Services of the Federal Government of Somalia. It provides free medical care, especially for inpatients. During the COVID-19 pandemic, the hospital exclusively served COVID-19 patients. The hospital has 115 beds and offers a range of services, including polyclinics, inpatient care, emergency services, and intensive care.

In 2019, there were a total of 61 health facilities run by public in the Benadir region, which encompasses the capital city [13]. De Martino hospital in this region serves as a secondary referral hospital (RH) indicating its role in providing specialized care. It’s important to note that while RHs like De Martino Hospital offer valuable specialist services, they may face challenges in meeting the extensive demand due to limited capacity and resources [14]. During the COVID-19 pandemic, De Martino Hospital emerged as a key center for managing COVID-19 cases, playing a vital role in the national response. This underscores its importance in addressing public health challenges, even during unprecedented events like a global pandemic.

### The study population and sample size

The study population is 210 HCWs working at De Martino Hospital between 5 August and 5 November 2022. HCWs include all hospital staff who provide health services directly or indirectly, such as managers, secretaries, doctors, nurses, laboratory technicians, radiologists, pharmacists, cleaners, security, or other personnel. All HCWs working in the hospital were aimed to be included in the study. In total, 204 HCWs were reached during data collection in the study and their informed consent was obtained using a written form.

### The study variables

To assess the HCWs’ compliance with COVID-19 prevention and control practices (IPC), the use of personal practices during their healthcare interactions in the hospital was measured using a self-reported questionnaire. Accordingly, the questionnaire form was created under three main headings; Personal Protective Equipment (PPE), Hand washing and Hygiene (HH) and other COVID-19 IPC practices. The data on personal characteristics, related factors to IPC, work conditions and practices on COVID-19 were also included in the questionnaire form. The questionnaire was adapted from the WHO’s risk assessment tool for healthcare workers in the context of COVID-19 [15], with some modifications according to the suitability of the hospital facility and using literature guidelines. The questionnaire is attached as “S1 File”.

The anonymity and confidentiality of the participants were ensured; a specific number was given to every questionnaire, there were no any information that could identify individual participants during or after data collection. A pilot study was conducted initially, then the questionnaire was distributed to the hospital staff and the data was collected between 5 August and 5 November 2022. There were 25 questions measuring the compliance of HCWs with COVİD-19; for PPE 12 questions, for HH 7 questions and for other COVID-19 IPC 6 questions IPC on the questionnaire. Responses to each question were measured using a 5-point Likert; “always (5 points), often (4 points), sometimes (3 points), rarely (2 points), or never (1 point)”. The scores of each person’s responses were categorized into two categories and the answers “always” and “most of the time” were taken as compliance. Then, the compliance responses of each participant were summed and the median was taken into account as the cut-off point; scores above the median were considered “high compliance”.

### Data analysis

For descriptive analyses, the variables measured by scale values were converted into categorical ones, and all independent variables were presented as percentages. In order to determine the association between independent variables and high compliance with the COVID-19 IPC measures, the Chi-square test was used by converting the scores of compliance to the COVID-19 IPC measures into two categories, and the answers “always” and “most of the time” were taken as “compliance”. Then, the compliance responses of each participant were collected and the median was considered as the cut-off point. Scores above the median were considered “high compliance” and presented by percentage for descriptive analysis.

In order to identify the key independent factors associated with a high level of compliance with COVID-19 IPC measures, variables that showed significant associations with high compliance in univariate analyses were included in logistic regression models. The backward LR method was used to identify predictive variables associated with the compliance of IPC measures and the association was presented with Odds ratio and 95% confidence interval.

Statistical Package for Social Sciences (SPSS 23) program was used in data analysis. P values lower than 0.05 were considered significant.

### Ethics approval

The study received ethical approval from Dokuz Eylul University’s Non-Interventional Research Ethics Committee (approval date: 17.08.2022, decision number: 2022/26-08) and adhered to the principles of the Declaration of Helsinki and local institutional guidelines. Official permission was granted by De Martino Public Hospital in Mogadishu, Somalia. Research participants were informed about the study’s purpose and methodology, and their consent was obtained during the data collection process.

## Results

In total, data from 204 HCWs were analyzed, achieving a response rate of 97%. Among the participants, 51% were male (Table 1, column 4). The majority of HCWs (46.1%) fell in the 20 to 29 age group, while only 15.7% were aged 40 and above (Table 1, column 5). Approximately, 77.8% held bachelor’s degrees or higher qualifications (Table 1, column 6). Nurses and paramedical staff constituted the largest group (51%), followed by (31.9%) doctors and doctor assistants (Table 2, column 4). Outpatient workers accounted for 20.1% of the participants, while inpatient workers made up 26.5%. HCWs in other clinical departments comprised 29.9% of the total (Table 2, column 5).

**Table 1.**
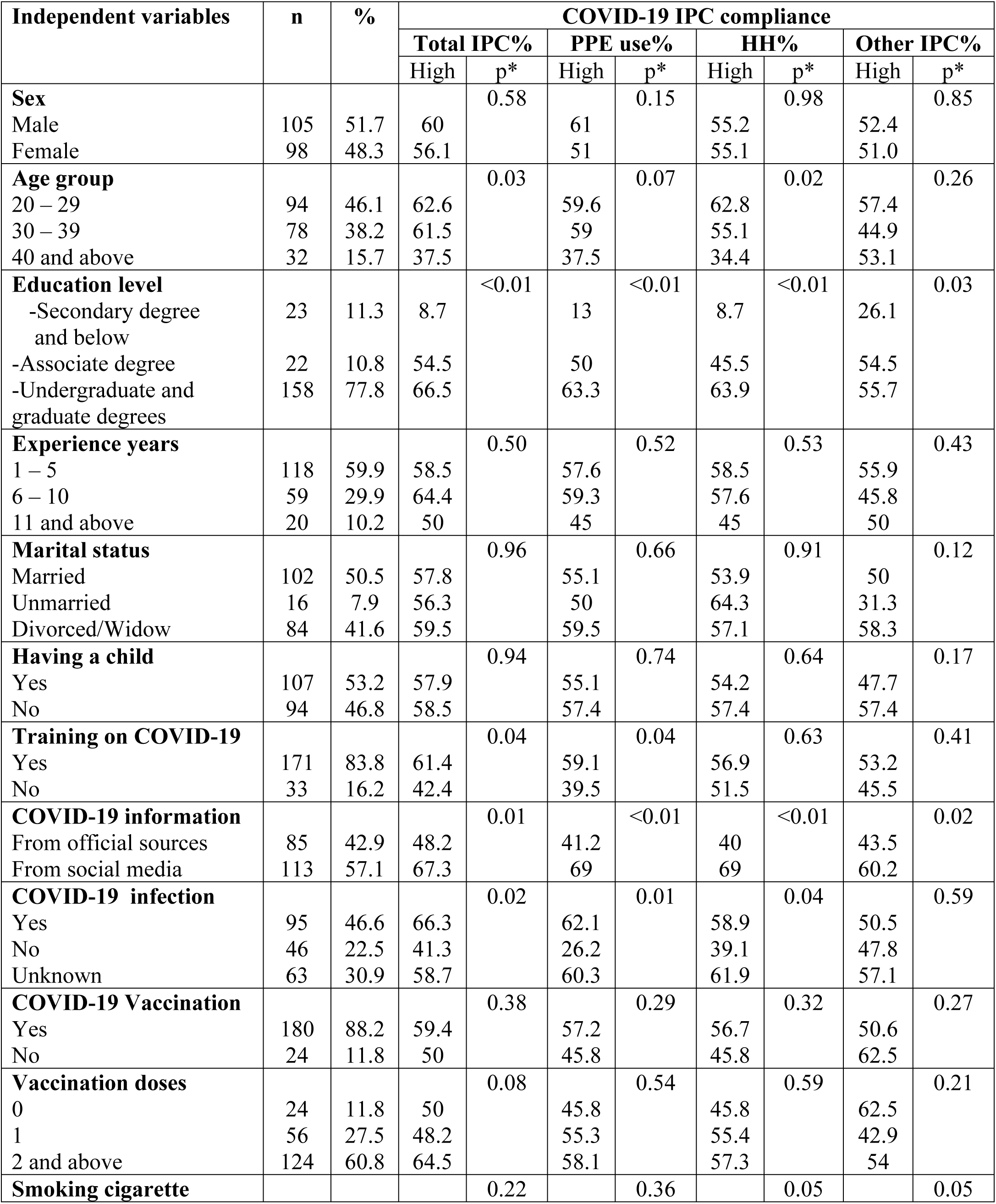
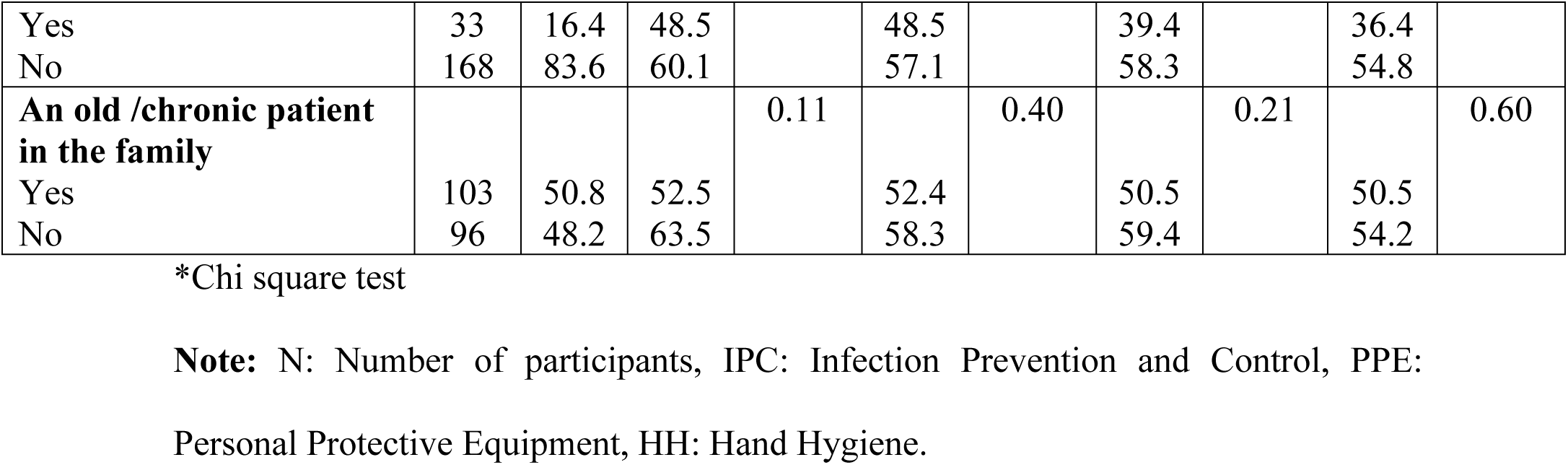
COVID-19 IPC compliance in HCWs during healthcare provision by sociodemographic characteristics and related factors to IPC.

**Table 2.**
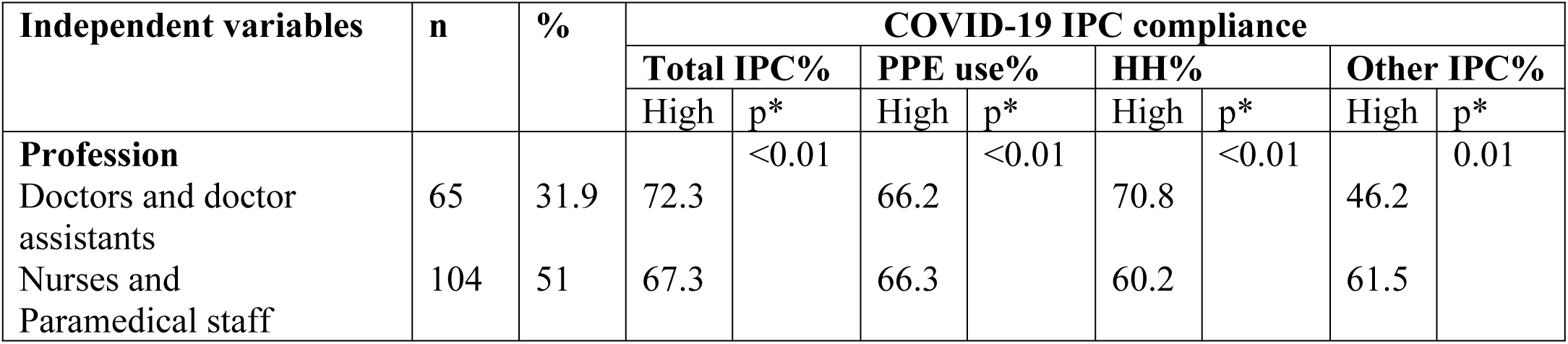
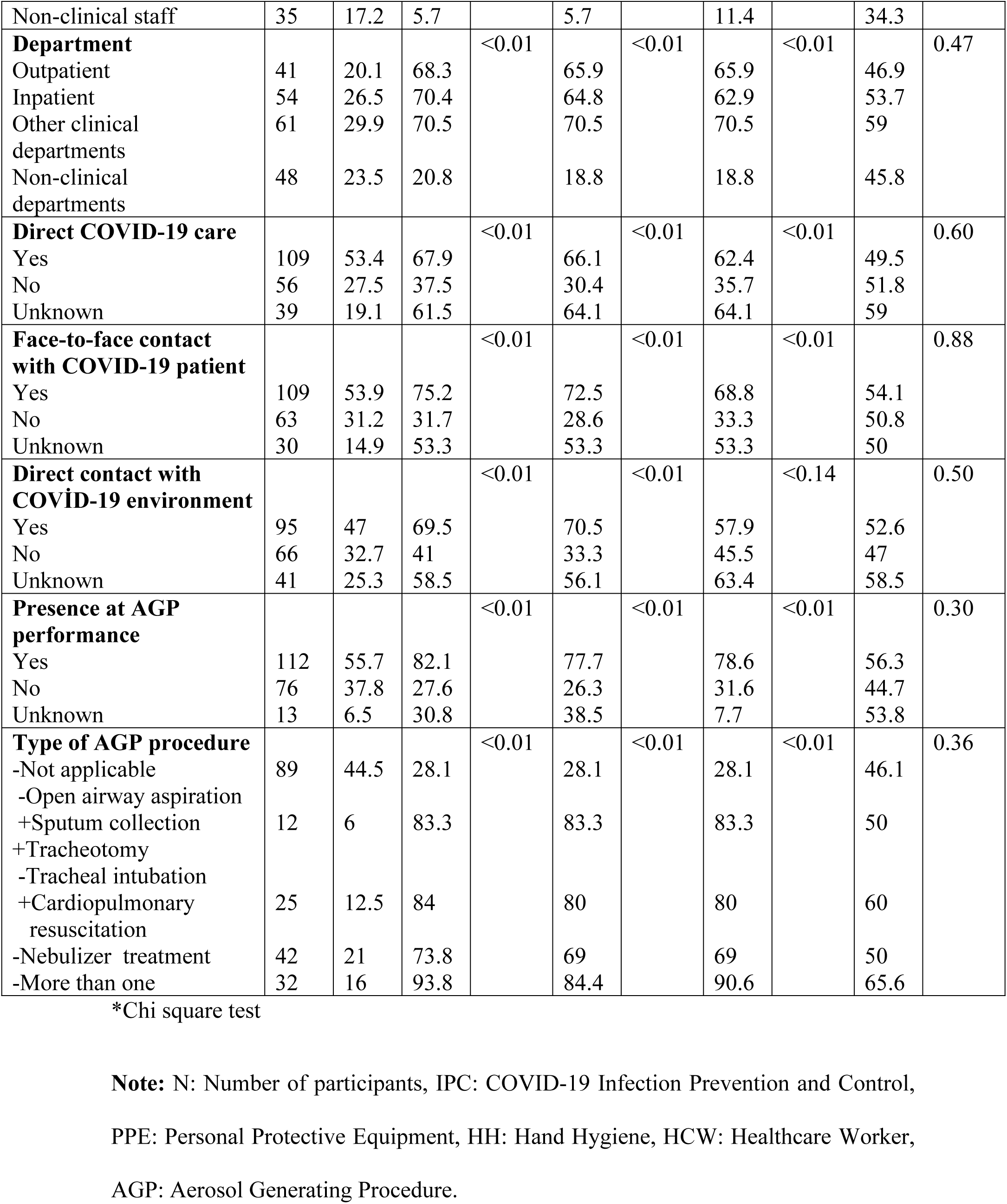
COVID-19 IPC compliance in HCWs during healthcare provision by working conditions and practices on COVID-19.

### Compliance of COVID-19 IPC practices among healthcare workers

The total IPC compliance was 58.3% among the participants. HCWs demonstrated 55.9% compliance with PPE during healthcare interactions, 55.4% for hand hygiene (HH), and 52% for other COVID-19 IPC measures during healthcare interactions (Fig 1).

**Fig 1.**
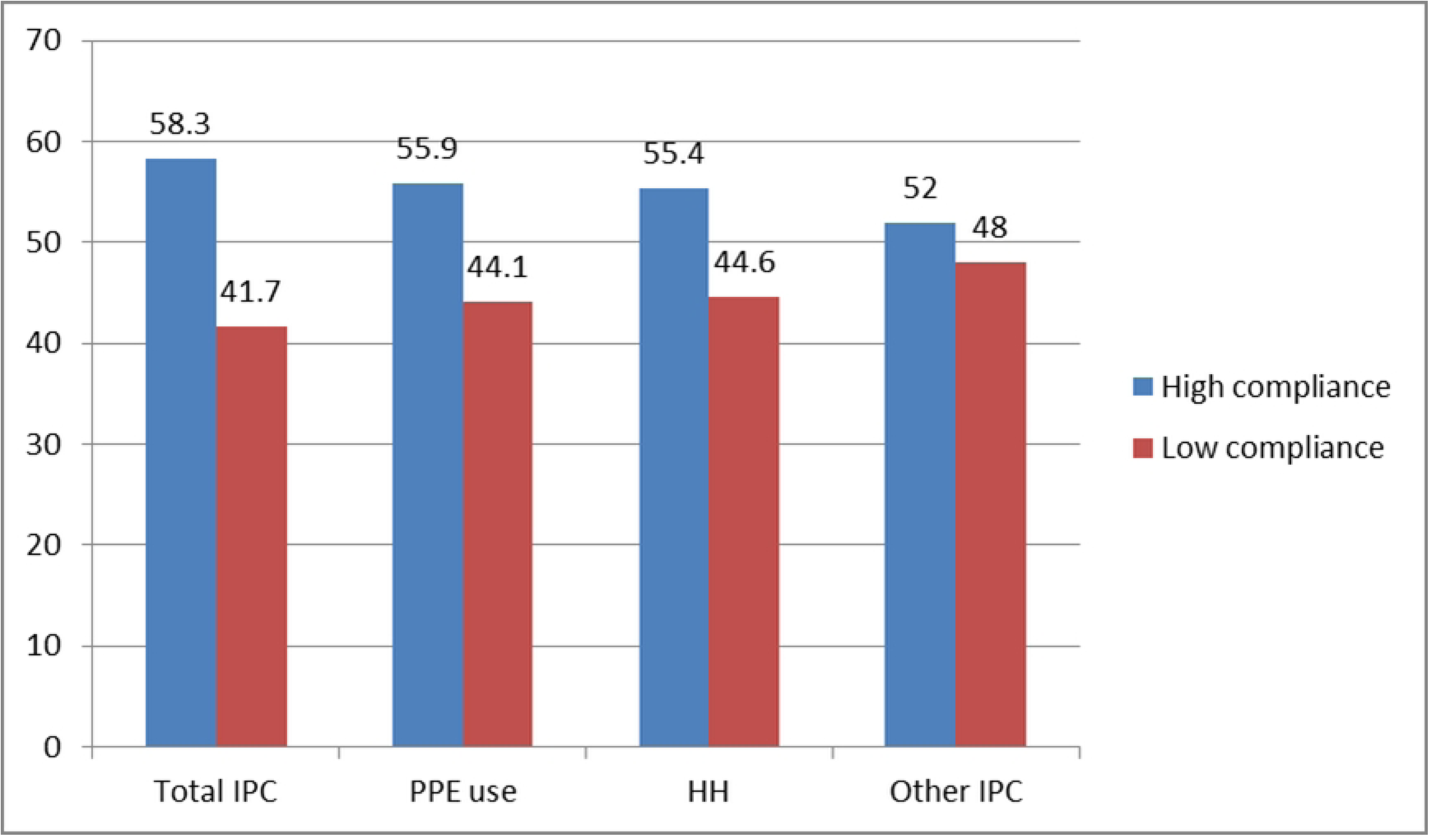
COVID-19 IPC compliance in the HCWs (Total IPC, PPE Use, HH, other IPC), %. **Note:** IPC: Infection Prevention and Control, PPE: Personal Protective Equipment, HH: Hand Hygiene.

Age and educational status were significantly associated with overall COVID-19 IPC and HH compliance (P<0.05 for all). Educational status was associated with compliance with PPE (p<0.01) and other COVID-19 IPC measures (p=0.03). Younger age groups and higher education level groups expressed better compliance (Table 1). There was no association between any of the IPC, PPE, HH compliance and marital status, having a child at home, having an old/chronic disease patient at home, or having COVID-19 vaccination. There was no significant association between the IPC measure dimensions and work experience years (Table 1). Participants who received training on COVID-19 reported higher compliance with total IPC and PPE than those who did not receive training (p<0.05).

There was a significant association between the type of HCWs and overall compliance with COVID-19 IPC, PPE and HH; nonclinical staff showed significantly lower levels of compliance for all three dimensions (p<0.01 for all). The same pattern was observed in the departments where HCWs worked; non-clinical departments showed lower compliance (p<0.01 for all). HCWs not providing direct care to COVID-19 patients, not having face-to-face contact, and not being present during the AGP performance reported lower compliance (p<0.01 for all). HCWs not having direct contact with the COVID-19 environment reported lower compliance in total IPC and PPE use (p<0.01 for all). There was no significant association with any of the working conditions and practices on COVID-19 and compliance with other IPC measures except the type of HCW (p=0.01) (Table 2).

In the logistic regression model, profession, having training on COVID-19, providing direct care to COVID-19 patients and presence when performing AGP were independent factors associated with total IPC compliance in HCWs. Doctors and doctor assistants and nurses and paramedical staff expressed a higher level of compliance with overall IPC compared to non-clinical staff (Doctors OR: 12.11, 95% CI: 2.23 – 65.84, nurses and paramedical staff OR: 21.38, 95% CI: 4.23 – 108.01). HCWs who received training on COVID-19 exhibited higher COVID-19 compliance than those who did not (OR: 3.48, 95% CI: 1.06 – 11.35). HCWs who were present when AGPs were performed exhibited higher compliance compared to those uncertain about that (OR: 12.45, 95% CI: 12.16 – 71.76) (Table 3).

**Table 3.** Independent determinants of COVID-19 IPC compliance in HCWs during healthcare provision: Results of multivariate logistic regression analysis* (Backward elimination method)

|  | **COVID-19 IPC compliance** |  |  |  |
| --- | --- | --- | --- | --- |
| **Variables in the model** | **Total IPC**<br>**OR (95% CI)** | **PPE use**<br>**OR (95% CI)** | **HH**<br>**OR (95% CI)** | **Other IPC**<br>**OR (95% CI)** |
| **Profession** |  |  |  |  |
| Doctors & doctor assistants | 12.11 (2.23 – 65.84) | 11.59 (1.94 – 69.01) | - | 1.35 (0.56 – 3.24) |
| Nurses & paramedical staff | 21.38 (4.23 – 108.01) | 17.91 (3.22 – 99.64) | - | 2.59 (1.13 – 5.93) |
| Non-clinical staff | Ref. | Ref. | - | Ref. |
| **Department** |  |  |  |  |
| Inpatient | - | - | 4.97 (1.62 – 15.21) | - |
| Other clinical departments | - | - | 6.2 (2.1 – 18.35) | - |
| Outpatient | - | - | 4.21 (1.34 – 13.23) | - |
| Non-clinical departments | - | - | Ref. | - |
| **COVID-19 material** |  |  |  |  |
| From official sources | - | 0.38 (0.17 – 0.83) | 0.45 (0.22 – 0.92) | - |
| From social media | - | Ref. | Ref. | - |
| **Direct COVID-19 care** |  |  |  |  |
| Provide | 0.26 (0.07 – 0.98) | 1.02 (0.38 – 2.76) | - | - |
| Unknown | 0.9 (0.2 – 3.99) | 4.10 (1.12 – 15.03) | - | - |
| Does not provide | Ref. | Ref. | - | - |
| **Face-to-face contact with COVID-19 patient** |  |  |  |  |
| Have | 1.5 (0.27 – 8.07) | - | 1.06 (0.32 – 3.51) | - |
| Does not have | 0.37 (0.07 – 1.98) | - | 0.4 (0.12 – 1.33) | - |
| Unknown | Ref. | - | Ref. | - |
| **Direct contact with COVID-19 environment** |  |  |  |  |
| Have | - | 2.59 (0.88 – 7.6) | - | - |
| Does not have | - | 1.08 (0.36 – 3.23) | - | - |
| Unknown | - | Ref. | - | - |
| **Presence at AGP performance** |  |  |  |  |
| Present | 12.45 (2.16 – 71.76) | 4.36 (0.8 – 23.79) | - | - |
| Not present | 1.59 (0.3 – 8.21) | 0.7 (0.13 – 3.52) | - | - |
| Unknown | Ref. | Ref. | - | - |
| **Training on COVID-19** |  |  |  |  |
| Received | 3.48 (1.06 – 11.35) | 2.66 (0.93 – 7.66) | - | - |
| Did not receive | Ref. | Ref. | - | - |
| **Education level** |  |  |  |  |
| Undergraduate and graduate degrees | - | - | 8.48 (1.64 – 43.94) | - |
| Associate degree | - | - | 3.67 (0.56 – 24.04) | - |
| Secondary degree and below | - | - | Ref. | - |

\*Variables included in the logistic regression model

For PPE; profession, the source of COVID-19 material and providing direct care to COVID-19 patient were independent factors. Compliance with PPE showed significant differences among HCWs in various roles. Doctors and doctor assistants (OR: 11.59, 95% CI: 1.94 – 69.01) and nurses and paramedical staff (OR: 17.91, 95% CI: 3.22 – 99.64) exhibited higher compliance compared to non-clinical staff. HCWs who accessed COVID-19 information from official sources expressed lower compliance compared to those who accessed information from social media (OR: 0.38, 95% CI: 0.17 – 0.83) (Table 3).

For HH, departments where HCWs worked, source of COVID-19 material and education level were independent factors. Clinical departments showed higher compliance compared to non-clinical departments (inpatient OR: 4.97, %95 CI: 1.62 – 15.21, outpatient OR: 4.21, %95 GA: 1.34 – 13.23, other clinical departments OR: 6.2, %95 GA: 2.1 – 18.35). HCW participants who read COVID-19 related material from Official sources reported lower compliance than those who read from social media (OR: 0.45, 95% CI: 0.22 – 0.92) (Table 3).

For Other IPC compliance, profession was the only predictor. Nurses and paramedical staff expressed higher compliance compared to non-clinical staff (OR: 2.59, 95% CI: 1.13 – 5.93) (Table 3).

For total COVID-19 IPC: age group, education, received training, source of COVID-19 information, covid infection, type of HCW, type of department, providing direct COVID-19 care, Having face-to-face contact with COVID-19 patients, having direct contact with the COVID-19 environment, presence at AGP performance and type of AGP.

For PPE: education, training, source of COVID-19 information, covid infection, Type of HCW, Type of department, providing direct COVID-19 care, having face-to-face contact with COVID-19 patients, having direct contact with the COVID-19 environment, presence at AGP performance and type of AGP.

For HH: age group, education, source of COVID-19 information, COVID-19 infection, type of HCW, type of department, providing direct COVID-19 care, having face-to-face contact with COVID-19 patients.

For Other COVID-19 IPC: education, source of COVID-19 information, Type of HCW.

## Discussion

This study was designed to assess COVID-19 IPC compliance among HCWs during healthcare interventions and determine the factors affecting their compliance. The study showed that 58.3% of HCWs had high compliance with COVID-19 IPC measures during healthcare interventions. The compliance was over 50% in all domains (PPE use, HH, and other IPC). Non-clinical staff had lower compliance compared with clinical staff .

There are several studies that reported high compliance with COVID-19 IPC in HCWs. In Ghana, a study with 424 HCWs in COVID-19 treatment centers reported high compliance with hand hygiene (88.4%) and PPE usage (90.6%) [16]. Two Ethiopian studies, involving 403 and 422 participants, found good COVID-19 infection prevention practices in 64.3% and 63.5% of healthcare workers, respectively. The first study revealed 96.1% compliance with hand hygiene but only 45.2% with PPE usage, possibly due to PPE availability, comfort, negligence, or education [17, 18].

Compared to previous studies, our findings indicated lower IPC compliance rates. This variance may be attributed to differences in study methods and the timing of data collection. While our study used compliance scores above the median for each domain, the referenced studies used either above-average scores or cutoff points of 60% or 75% of total compliance scores. It is also possible that healthcare workers’ adherence to IPC measures decreased over time since the pandemic’s onset. A study on healthcare worker HH practices observed a 13.7% increase upon room exit during the initial COVID-19 wave. Compliance dropped by 9.9% post-lockdown but rebounded by 2.8% in the second wave [19].

Some studies have reported low HCW compliance with COVID-19 IPC measures. For instance, in a study involving 422 HCWs at COVID-19 referral hospitals in Ethiopia, overall compliance with COVID-19 prevention practices was only 22% [20]. In this study, only 63.4% of participating HCWs received training on COVID-19 and 58.2% read COVID-19 materials and 83.2% of HCWs felt a shortage of appropriate PPE in the hospital.

A review identified various barriers to HCWs’ compliance with IPC guidelines for respiratory infectious diseases, including the availability of training programs, PPE supply, and individual factors such as knowledge, attitude, beliefs, and PPE discomfort [21]. Additionally, a study conducted in Uganda, involving 657 HCWs at community hospitals, found that only 37.0% of participants had good COVID-19 IPC practices, despite high rates of mask usage and hand washing [22].

In our study, doctors, doctor assistants, nurses, and paramedical staff demonstrated higher COVID-19 IPC compliance than non-clinical staff, except in hand hygiene (HH) practices. This aligns with prior research. For instance, a study performed in Private-Not-for-Profit community hospitals in Uganda found a significant association between clinical HCWs and good COVID-19 IPC practices [22]. Similarly, a study in COVID-19 treatment centers in Ghana revealed that non-clinical staff exhibited significantly lower compliance with hand hygiene and PPE usage compared to clinical staff [16].

The difference in compliance may be due to the higher risk faced by clinical healthcare workers in close contact with COVID-19 patients. A study from Somalia, reported that ancillary staff, including security workers and cleaners, have a higher infection risk due to lower knowledge and adherence to infection control measures when handling suspected COVID-19 patients. Healthcare assistants are often informally employed and receive less attention than formal employees like doctors, nurses, and technologists [11].

Our study showed that HCWs who were present during AGPs exhibited higher compliance with overall IPC measures compared to those uncertain about that. This aligns with Ashinyo ME et al.’s study that found high compliance with COVID-19 IPC protocols during AGPs [16]. Healthcare workers performing AGPs face a higher risk of COVID-19 infection, possibly explaining their heightened compliance [23].

Our study also showed that HCWs who had received training on COVID-19 exhibited higher COVID-19 compliance with overall IPC than those who did not. This is in line with 2 studies on 422 HCWs in Ethiopia; Etafa W. et al [20] and Arsemahagn MA (24), and a review study by Cooper S. et al. [25].

Interestingly, our study observed that HCWs who obtained COVID-19 information from official sources showed lower compliance with PPE and HH compared to those who accessed information from social media. In contrast, a study on the Somali population reported the opposite, where HCWs who relied on social media for COVID-19 information exhibited lower compliance, likely due to misinformation [26]. However, De Martino HCWs may follow specific social media pages they trust for COVID-19 information.

To improve compliance, HCWs must receive continuous awareness and training in COVID-19 IPC guidelines. Policymakers should develop comprehensive programs to raise awareness among HCWs at all levels and provide the necessary equipment and supplies for effective IPC practices in healthcare settings.

The study has limitations. Firstly, there could be recall bias; because participants were asked about their compliance with COVID-19 IPC measures during the late stages of the pandemic when no COVID-19 patients were likely admitted, and some PPE like respirators, gowns, and face shields might not have been used. To minimize this bias, we asked about daily IPC practices and included specific questions for suspected or confirmed COVID-19 patients. Since the study was a single-centre study, De Martino Public Hospital may not fully represent other pandemic hospitals in Somalia, although it plays a significant role in treating COVID-19 patients. Nonetheless, the inclusion of all staff members and high response rate enhances the generalizability of the results within the hospital. Furthermore, the limited number of participants has resulted in a wide confidence interval for the odds ratio derived from the model.

## Conclusions

A notable portion of hospital workers reported inadequate compliance with COVID-19 IPC measures, with particularly low adherence among non-clinical staff. This lower compliance might be linked to their perception of low risk and a lack of awareness regarding to COVID-19. It is essential to recognize that all HCWs in the hospital are susceptible to COVID-19 infection. Therefore, prioritizing awareness campaigns and behavior change interventions, especially among non-clinical staff, is crucial for effective COVID-19 infection prevention and control within the hospital.

## Data Availability

All relevant data are within the manuscript and its Supporting Information files

## Acknowledgments

First; we give our sincere gratitude to the managers of De Martino hospital for allowing us to conduct our study on the hospital.

Second; we thank Dr. Lul Ahmed Abdi, Maternity department, De Martino hospital who was the link person between us and the hospital administration. Also she supported us in the delivering of the questionnaire papers to the hospital workers and collecting it after collection during the data collection process, since she works at the hospital and knows the hospital well.

Third; we extend our gratitude to the different groups of the healthcare workers at De Martino hospital for participating in the study.

## Supporting information

**S1 File: Questionnaire for compliance of healthcare workers with COVID-19 prevention and control measures**

**S1 Table. Sociodemographic characteristics, related factors to infection prevention and control, working conditions and practices on COVID-19 of the healthcare workers**

**S2 Table. Healthcare workers’ COVID-19 infection prevention and control measure dimensions (Personal protective equipment use, Hand Hygiene and other infection prevention and control measures)**

